# RTBES Radiological Index for Quantitative Assessment of Tuberculosis Evolution: Development and Validation

**DOI:** 10.1101/2025.02.14.25322286

**Authors:** Patrícia Cuadras, Gerard Rafart, Martí Català, Lilibeth Arias, Ricard Pérez, Neil Martinson, Alex Rosenthal, Andrei Gabrielian, Sergo Vashakidze, Montserrat Tenesa, Jordi Bechini, Cristina Vilaplana

## Abstract

**Question addressed by the study:** A radiological index capable of quantifying pulmonary tuberculosis (TB) disease progression on standard Chest X-Ray (CXR) could enhance clinical management and inform therapeutic decision-making. Our objectives were: to develop a radiological index that quantitatively assesses the progression of pulmonary TB in patients over the course of treatment and follow-up, to evaluate its usefulness by using CXRs from a cohort of TB patients, and to validate it with CXRs from an independent cohort of TB patients.

**Materials/patients and methods:** An index was developed to identify and classify patterns of radiological findings of both active and inactive pulmonary TB. This index was evaluated in a first cohort comprising a pseudonymized sample of serial CXRs from 21 TB patients at the Perinatal HIV Research Unit (PHRU) in South Africa and validated in a second, larger, independent cohort comprising 50 treated TB patients from the National Institute of Allergy and Infectious Diseases (NIAID) TB Data Portals. The relationship between index values for activity markers and patients’ clinical characteristics was also analysed.

**Results:** A radiological index, the RUTI-TB Evolution Score (RTBES), was created and validated in two cohorts, achieving 95.96% interobserver agreement (±2 points). In both cohorts, RTBES for active TB signs decreased over follow-up. Higher baseline RTBES correlated with male sex, higher bacterial loads, and extensive pulmonary involvement (including cavitation and pleural effusions). Baseline RTBES≥2 predicted unfavourable outcomes; 80% of failures/deaths occurred above this threshold.

**Answer to the question:** RTBES effectively quantified pulmonary TB severity and tracked treatment evolution on standard CXRs, showing promise as a prognostic tool to inform clinical decision-making.

## 1. Introduction

The Chest X-Ray (CXR) is the most frequently performed imaging study. It is relatively affordable, and portable equipment allows it to be taken in nearly any setting. CXR continues to be the first radiological exploration for assessments of possible pulmonary pathology. A systematic, rigorous, precise, and thorough approach is essential when evaluating CXRs. Although all radiological modalities have technical limitations, advances in digital radiology have reduced many of the traditional limitations of CXRs [1]. The introduction of technologically improved, lightweight, portable devices with battery and internet connectivity has simplified their application across diverse environments.

The utility of CXR in diagnosing and monitoring pulmonary tuberculosis (TB) is well-established in clinical practice, though its sensitivity and specificity is often viewed as insufficient. However, over the past decade, various TB prevalence studies have demonstrated that CXR remains the most sensitive diagnostic tool for pulmonary TB screening [2]. As a result, CXR has regained prominence in diagnostic and follow-up (F-U) algorithms for pulmonary TB. There is an increased need for systems that quantify radiological findings in pulmonary involvement, enabling standardized comparisons between patients and in the same patients during treatment. These systems, referred to as indices, assign numerical weights to each specific CXR variables related to pulmonary TB. Since the 1960s, various radiological indices have been developed for assessing pulmonary TB in clinical trials and for TB screening across different patient populations [3–12].

Existing indices have also been assessed as screening tools in populations with low HIV prevalence or used in hospital settings for the initial management of patients suspected of having pulmonary TB. However, these indices frequently exhibit significant intra- and inter-observer variability, limited sensitivity, and lack validation across diverse populations and settings, which restricts their universal applicability in epidemiological studies, in populations with varying HIV prevalence, and among both outpatients and hospitalized patients.

Radiological indices custom-designed for pulmonary TB typically combine clinical and/or microbiological findings with radiological findings. These indices are often complex, involving qualitative, semi-quantitative, or cumbersome quantitative parameters. Although existing indices have been evaluated for intra- and inter-observer agreement, diagnostic accuracy, and reproducibility, simple, quick, and easy-to-use indices nevertheless are still lacking. Existing indices for quantifying pulmonary TB disease underscore the need to develop a purely radiological index with enhanced diagnostic precision Such an index should provide simple and reliable quantification of parenchymal, pleural, and thoracic lymphadenopathy patterns in active pulmonary TB and its sequelae, with minimal inter-observer variability across different populations. A new CXR index with this capacity could be highly useful in clinical practice, aiding in pulmonary TB diagnosis, tracking disease progression, and predicting treatment outcomes to identify patients at high risk of treatment failure or mortality, ultimately improving therapeutic strategies.

Based on this rationale, we hypothesize that a radiological index capable of quantifying the degree of pulmonary TB disease progression on standard CXR could enhance clinical management and inform therapeutic decision-making. Our objectives were: firstly, to develop a radiological index that quantitatively assesses the progression of pulmonary TB in patients over the course of treatment and F-U; secondly, to evaluate its usefulness by using CXRs from a cohort of TB patients; and thirdly, to validate the score with CXRs from an independent cohort of TB patients.

## 2. Methods

### 2.1. Index Development

The following principles guided the development of the index: simplicity and ease of use, allowing for application in clinical contexts by non-radiologists and even non-medical personnel; it had to be reliable, with minimal inter-observer variability; it required accuracy in assessing the diverse presentations of pulmonary TB; it needed to quantify both active and inactive radiological signs of TB and reflect post-TB sequelae; results had to be expressible as numerical variables; the index needed to be easily adaptable for automated analysis software.

A preliminary version of the index was developed to identify and classify radiological patterns of acute TB involvement in the parenchyma (cavities, infiltrates, bronchogenic dissemination, miliary TB), lymph nodes (mediastinal, hilar), and pleura, as well as chronic or residual changes in these same areas, based on findings from both active and inactive pulmonary TB. Variable selection was done according to the consulted literature [4, 8, 12–24] and professional experience of the investigator team.

### 2.2 Evaluation of the index in Cohort 1

A pseudonymized sample of serial CXRs from a cohort of 21 TB patients at the Perinatal HIV Research Unit (PHRU) in South Africa was used for this study. The sample comprised 21 patients, including 8 women and 13 men. The CXRs, taken between 2013 and 2014, were evaluated in a blinded fashion by two radiologists (a faculty member and a resident). Patient ages, culture-negative conversion dates, HIV serology or other immunosuppression statuses, as well as survival and final patient status, were not available.

Approval was obtained from the Human Research Ethics Committee of the University of Witwatersrand in Johannesburg, South Africa, to use CXRs from patients included in the study early treatment response in smear-positive and smear-negative/GeneXpert positive adult TB patients with HIV co-infection, led by Dr N. Martinson.

CXRs of suboptimal quality in any technical aspect were excluded from the study. The quality criteria for CXRs included: patient and X-ray tube positioning: no rotation, and images should not be lordotic; degree of inspiration: visibility of 9-10 posterior rib arches or 6-8 anterior rib arches; absence of patient movement; appropriate mA and kV settings, with adequate contrast. For 16 patients, three F-U CXRs were assessed, and for 5 patients, two F-U CXRs were assessed. In total, 58 posteroanterior (PA) CXRs were evaluated in the upright position, starting from diagnosis through subsequent F-Us (Figure 1).

**Figure 1.**
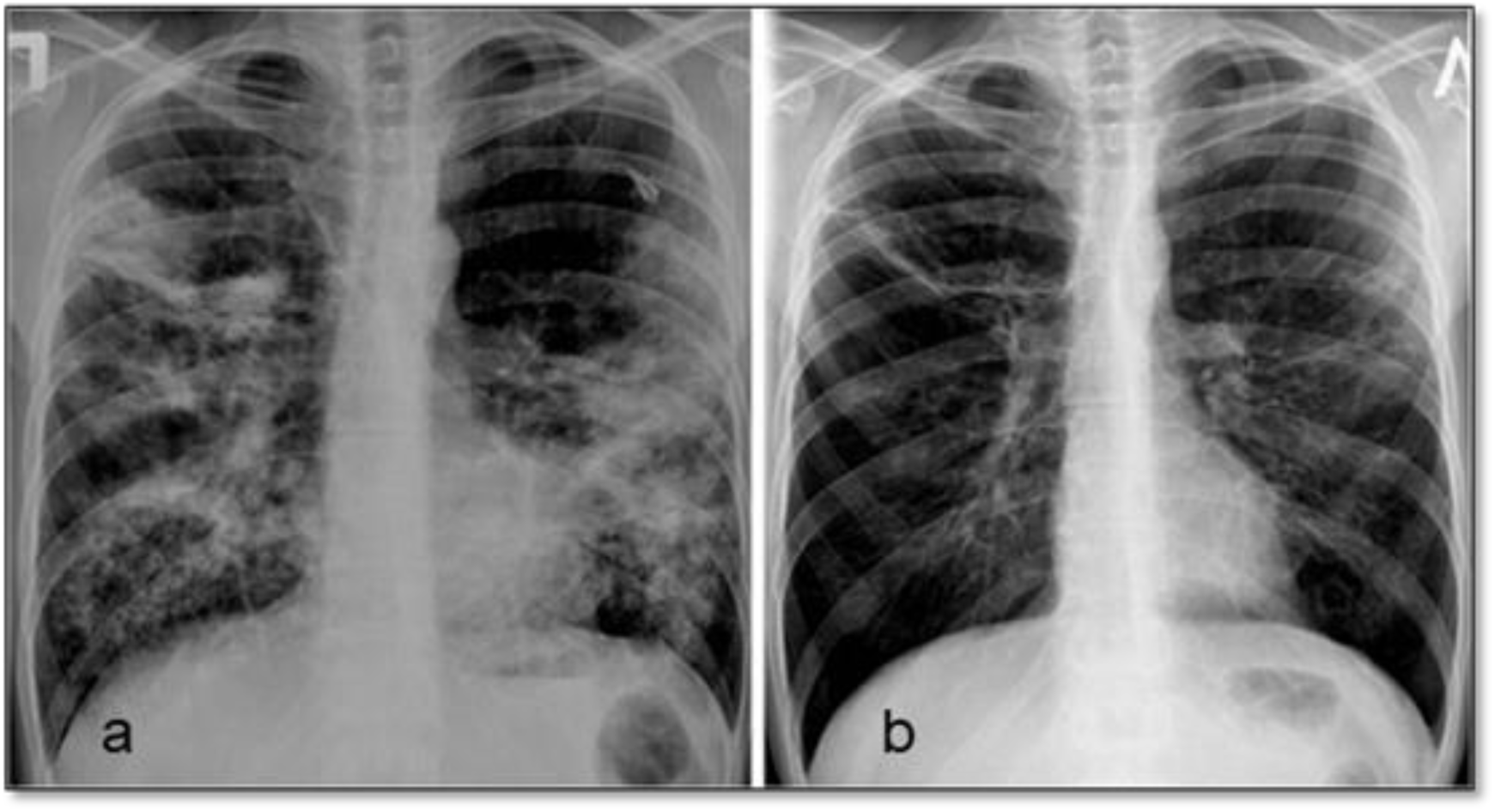
P-A CXR with bilateral consolidations. Panel a is a BL CXR and Panel B is a CXR done during a FU.

Based on this first evaluation, the initial index was updated, and this final version (RTBES score, Supplementary information) was validated in an independent cohort.

### 2.2. Validation in an Independent Cohort

We then validated the index using CXRs from a second, larger, independent cohort of TB patients and assessed the relationship between index values for activity markers and patients’ clinical characteristics. The recording of inactive markers in this study allowed for objective verification of their stability over time; however, these were not used in the index validation. A pseudonymized sample of serial CXRs from 50 treated individuals with TB, provided by the National Institute of Allergy and Infectious Diseases (NIAID) through a publicly available database, the NIAID TB Data Portals (https://tbportals.niaid.nih.gov/), was analysed. The cohort included 20 women (ages 19–57) and 30 men (ages 23–84) from Belarus. Clinical and microbiological data were available, and each patient had between 5 and 17 CXRs. In total, 365 CXRs, taken retrospectively from April 15, 2010, to May 11, 2015, were evaluated. Radiological assessment was performed independently by two observers, blinded to all clinical and microbiological data.

### 2.3. Analysis of results according to clinical characteristics

For this we focused on the evolution of the index in active TB signs during the first twelve months post-diagnosis and treatment initiation. Evaluation of inactive signs was omitted, as these remained stable over time. Cohort 2 data and corresponding CXR records were included.

Given the variability in the timing of CXR and differences in the number of CXRs available per patient, we grouped the F-U points to standardize results presentation into four key timepoints. The baseline (BL) timepoint included the first CXR obtained at diagnosis. The first F-U point (FU1) aggregated CXRs performed in the second- or third-month post-diagnosis. The second F-U point (FU2) included CXRs from the fourth, fifth, or sixth month, while the third F-U point (FU3) combined CXRs taken in months ten through fourteen.

A table summarizes the specific CXRs used for each patient at these analysis timepoints. All fifty patients had CXRs available for BL. For FU1, thirty-six patients had CXRs; for FU2, thirty-two; and for FU3, thirty-two patients. Additionally, twenty-two patients had CXRs available for BL, FU1, and FU2, while twelve patients had CXRs available for all four timepoints (Table 8).

Differences in the overall index values at BL and at each F-U (FU1, FU2, FU3) were analysed in relation to clinical characteristics of the patients. For certain clinical characteristics, analyses were first conducted using all available CXRs, and subsequently using only those at timepoints BL, FU1, FU2, and FU3.

To determine the final patient outcome, final states were defined based on the current definitions established by the WHO [25]. Five possible outcomes were identified: cure, treatment completed, treatment failure, lost to follow-up, and death.

### 2.4. Statistical Methods

Data collection was conducted using Microsoft Excel®. The process involved an initial phase of database review and validation, performed for each variable of interest. For quantitative variables, mean and standard deviation were calculated. For qualitative variables, absolute and relative frequencies were used. Statistical analyses were implemented using the MATLAB® platform. An analysis of variance (ANOVA) was performed to compare the means and variances of continuous variables across different groups. Fisher’s exact test was used for comparing categorical variables across groups. A significance level of α=0.05 was selected.

The probability calculation for favourable or unfavourable outcomes was conducted using logistic regression, with the generalized linear model fitting function (glmfit). Inter-reader agreement for CXR assessments by the two radiologists was evaluated using the concordance index.

## 3. Results

### 3.1. Index developed

A first version of the index was developed based on activity and non-activity findings as per described in the section 2.1. The index developed was further refined with CXR from Cohort 1. Details on the final variables included and analysis methods are provided in the procedure developed ad-hoc (Supplementary Information S1). This final index was named RUTI-TB Evolution Score (RTBES).

### 3.2. Inter-Observer Agreement

To evaluate reproducibility and reliability, the agreement between the two blinded radiologists was assessed. Index values for TB activity signs were compared for each patient at each time point. In 82.89% of cases, the difference in total scores for active signs between the two observers ranged from 0 to 1 point. When the allowed difference was expanded to 2 points, the agreement increased to 95.96%.

### 3.3. RTBES is useful to quantify TB radiological progression

An initial evaluation of each patient’s score was conducted at BL and at each F-U for both active and inactive TB signs. The results indicated that the index for active TB signs decreased over time, while the index for inactive signs remained stable, thereby confirming the index’s capacity to measure disease dynamics. This observation was true for cohort 1 and for cohort 2, as well as when combining the results from both cohorts. Results of the mean index value at BL and the index variation over time both for each cohort and when combined is presented in Table 1.

**Table 1.** Mean Index Values for Signs of TB Activity and Inactivity at Baseline (BL) and during Follow-Up (F-U). Mean Index Values for Active and Non-active TB signs at BL and First F-U, Including Index Variation Between BL and the First Follow-up, and Between BL and the Last Available Follow-up, by Cohorts and Combined Cohorts. BL: Baseline; F-U: Follow-Up.

| CXR findings | BL Index Value | Index Value at the Last Available F-U | Variation in Index Value Between BL and the First F-U | Variation in Index Value Between BL and the Last Available F-U |
| --- | --- | --- | --- | --- |
| **Cohort 1** |  |  |  |  |
| Active TB signs | 3.33±3.32 | 2.13±4.5 | -0.70±2.62 | -1.20±5.39 |
| Non-active TB signs | 1.11±1.57 | 1.08±1.49 | -0.04±0.62 | -0.03±1.03 |
| **Cohort 2** |  |  |  |  |
| Active TB signs | 2.63±2.78 | 2.35±5.36 | +0.38±3.01 | -0.28±5.84 |
| Non-active TB signs | 1.34±1.71 | 1.53±1.70 | +0.07±0.82 | +0.19±1.10 |
| **Combined cohorts (1+2)** |  |  |  |  |
| Active TB signs | 3.04±3.12 | 2.22±4.89 | -0.26±2.83 | -0.82±5.58 |
| Non-active TB signs | 1.21±1.71 | 1.27±1.59 | 0.00±0.71 | +0.06±1.06 |

### 3.3. Results Analysis Based on Clinical Characteristics

Regarding the overall RTBES, no correlation was found between the score and smoking status, BMI, initial TB culture results, or TB type based on drug resistance profiles. However, a statistically significant correlation was observed between the overall index value and the patient’s gender, as well as the presence and quantity of Acid-Fast Bacilli detected by microscopy (Table 2).

**Table 2.** Index Values for Signs of Activity at Baseline (s0), According to Different Evaluated Clinical Characteristics. SD: Standard Deviation; N = Number of patients; AFB: Acid-Fast Bacilli; DS: Drug-Sensitive; DR: Drug-Resistant; MDR: Multi-Drug Resistant; XDR: Extensively Drug-Resistant. Statistically significant values are highlighted in bold (p ≤ 0.05).

| Clinical and microbiological characteristics | Mean ± Standard Deviation | p-value |
| --- | --- | --- |
| **Biological sex** |  |  |
| Females (N=20) | **1.7 ± 1.7** | **0.05** |
| Males (N=30) | **3.3 ± 3.2** | **0.05** |
| **Smoking habit** |  |  |
| Smoker (N=30) | 3.0 ± 3.0 | 0.28 |
| Non-smoker (N=20) | 2.1 ± 2.4 | 0.28 |
| **BMI** |  |  |
| Low weight (N=10) | 3.1 ± 2.8 | 0.51 |
| Normal (N=30) | 2.4 ± 2.7 | 0.45 |
| Preobesity (N=9) | 3.2 ± 3.3 | 0.53 |
| Obesity (N=1) | 0.0 ± 0.0 | 0.35 |
| **Sputum Microscopy (Presence/Absence of AFB)** |  |  |
| Negative (N=38) | **2.2 ± 2.6** | **0.04** |
| Positive (N=12) | **4.1 ± 3.0** | **0.04** |
| **Sputum Microscopy (AFB/ Field)** |  |  |
| Negative (N=38) | **2.2 ± 2.6** | **0.04** |
| 1-9/100 (N=6) | 4.2 ± 3.6 | 0.15 |
| 10- 99/100 (N=3) | 4.0 ± 1.7 | 0.38 |
| 1-9/1 (N=1) | 1.0 ± 0.0 | 0.56 |
| 10-99/1 (N=2) | 5.5 ± 3.5 | 0.14 |
| **Sputum culture (bacilli/mL)** |  |  |
| Negative (N=6) | 1.4 ± 2.0 | 0.26 |
| 1-19 (N=12) | 1.4 ± 1.4 | 0.08 |
| 20-100 (N=18) | 3.3 ± 3.1 | 0.20 |
| 100-200 (N=10) | 2.9 ± 3.2 | 0.78 |
| >200 (N=4) | 4.5 ± 3.3 | 0.16 |
| **Drug susceptibility** |  |  |
| DS-TB (N=13) | 2.3 ± 2.8 | 0.63 |
| Mono DR (N=3) | 1.7 ± 1.5 | 0.54 |
| Poli DR (N=2) | 3.0 ± 4.2 | 0.85 |
| MDR no XDR (N=16) | 2.2 ± 3.0 | 0.48 |
| XDR (N=16) | 3.4 ± 2.8 | 0.16 |

#### Pulmonary Involvement and Follow-up Analysis

Significant statistical differences were observed in the index values for TB activity signs. Patients with a higher number of pulmonary cavities, larger cavity sizes, pleural effusions, or concurrent pulmonary and extrapulmonary TB had higher activity index values (Table 3).

**Table 3.**
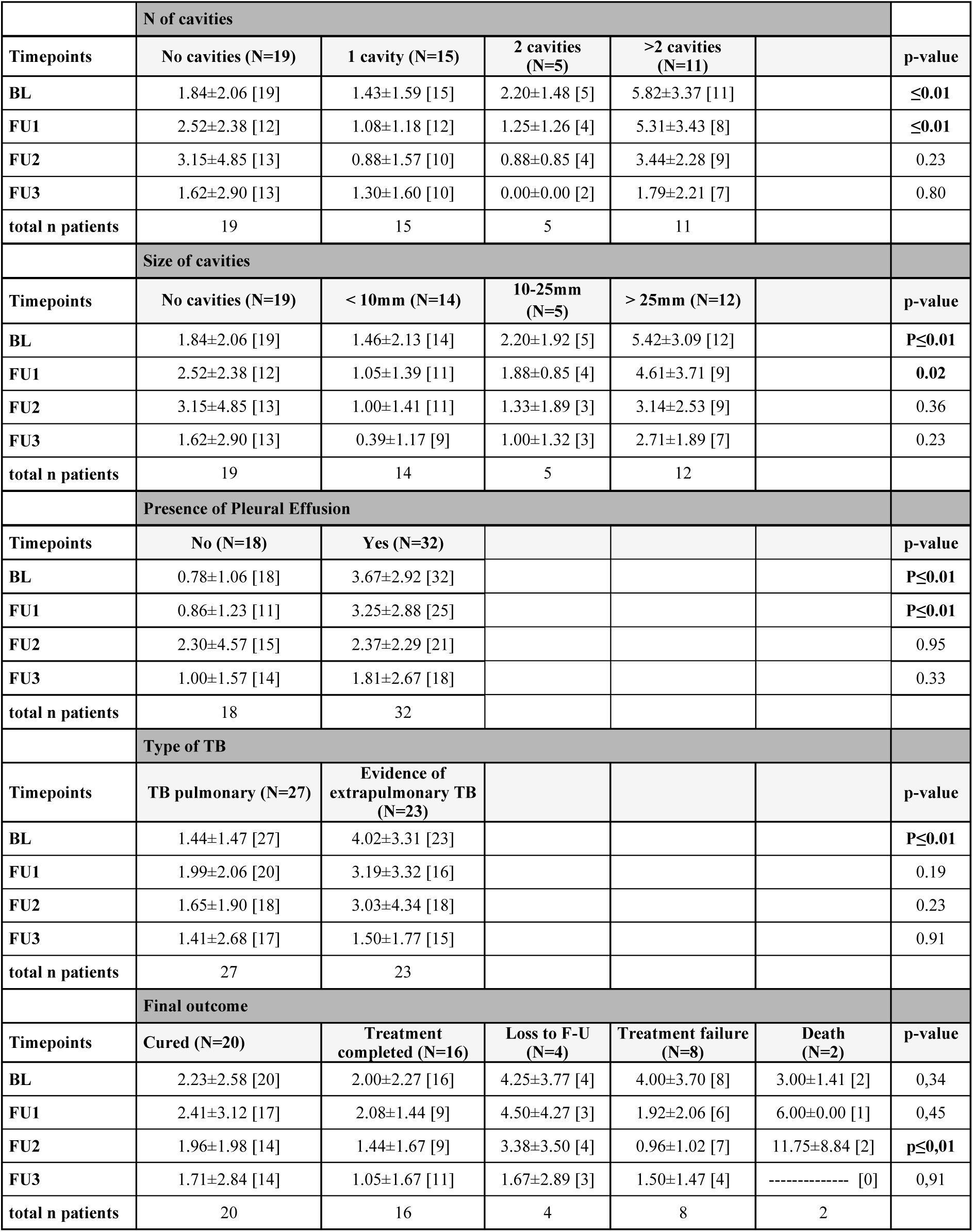
Statistical Significance of overall Index Values for TB Activity Signs, based on the different clinical characteristics over time. Results are presented as means ± Standard Deviation; N: Number of patients; [..]: Number of patients at each time point. Statistically significant values are highlighted in bold.

It was also noted that in these patients – with more extensive pulmonary, pleural, or systemic disease – the activity index decreased over time up to follow-up point s3. However, the decline did not reach the lower index values seen in patients with less severe pulmonary or pleural disease or without extrapulmonary dissemination. For each potential outcome of the evaluated variables, it was assessed whether the activity index values differed statistically between patients with that specific outcome and those with other outcomes for the same variable.

Patients with a final outcome of cure or treatment completion had a significantly lower index value compared to patients with other final outcomes.

#### RTBES according to the Patient’s Final Outcome and Follow-up Point Correlation between various clinical and radiological variables

When we analysed clinical variables, we observed that the sample was not homogeneous (Figure 2). Men were more likely to be smokers, to have a positive culture, and to exhibit a higher bacterial count per millilitre compared to women. Non-smokers tended to have negative cultures, and when cultures were positive, the bacterial count per millilitre was lower. Patients with positive sputum microscopy had a higher likelihood of a positive culture with counts exceeding 20 bacilli per field, and they were also more likely to have larger cavities. All patients with multidrug-resistant TB (MDR-TB) or extensively drug-resistant TB (XDR-TB) had positive cultures. Those with a greater number of cavities also tended to have larger cavities. Patients with pleural involvement were more likely to present with multiple cavities, positive sputum microscopy, and positive cultures with higher bacterial counts compared to those without pleural involvement. Patients with both pulmonary and extrapulmonary TB were more likely to have positive sputum microscopy, as well as positive cultures with higher bacterial counts. Additionally, individuals with a final outcome of treatment failure or death were more likely to have XDR-TB or MDR-TB. Finally, patients who experienced treatment failure were more likely to have both positive sputum microscopy and positive cultures.

**Figure 2.**
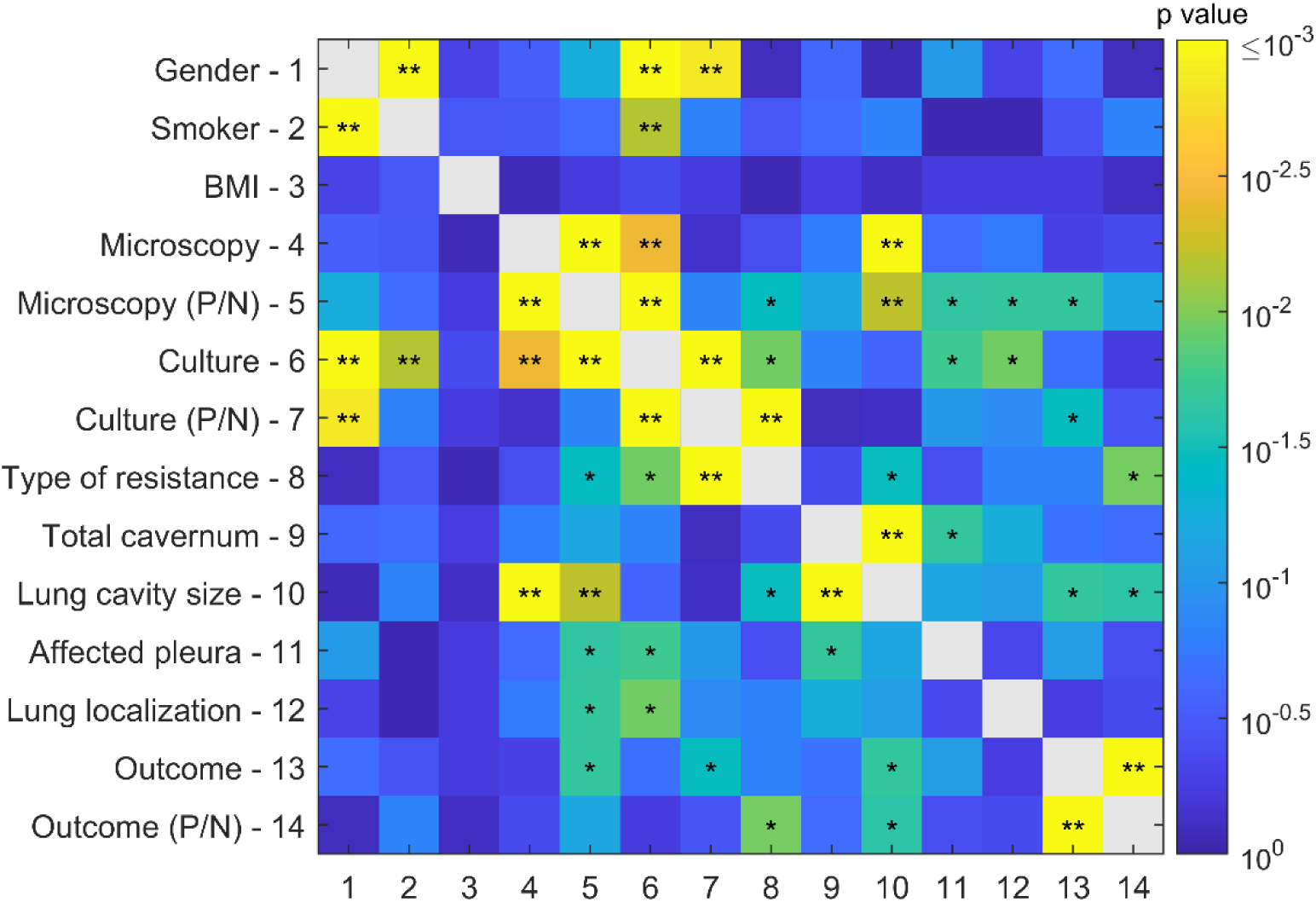
Relationship between the different clinical characteristics of the patients. *corresponds to 0.01 < p ≤ 0.05, and ** to p ≤ 0.01. P: positive; N: negative.

Then we observed that the activity index values at BL were significantly higher for patients presenting specific pairs of variables compared to those without, and that these pairs were also associated with a significantly higher frequency of treatment failure or death (Table 4).

**Table 4.**
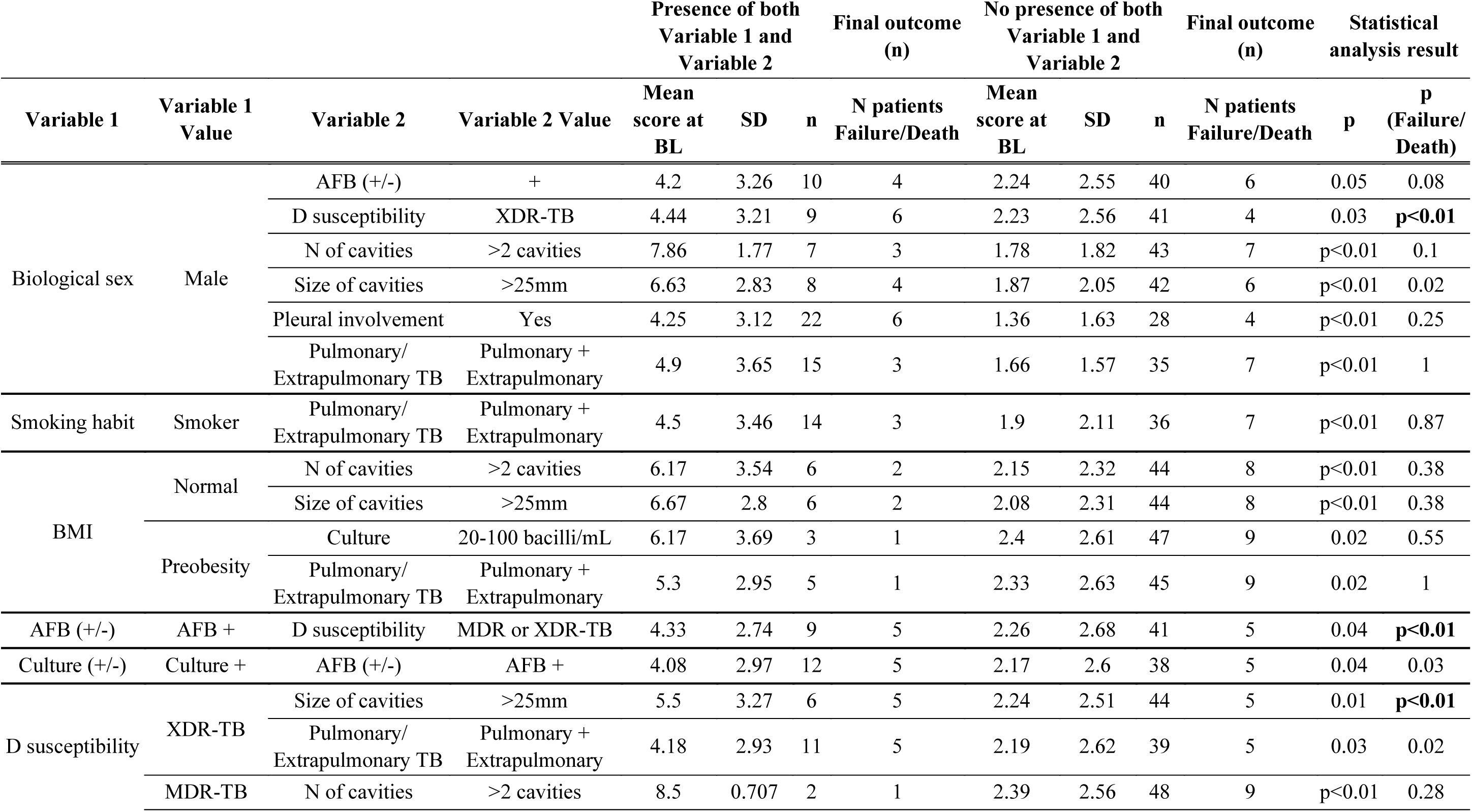
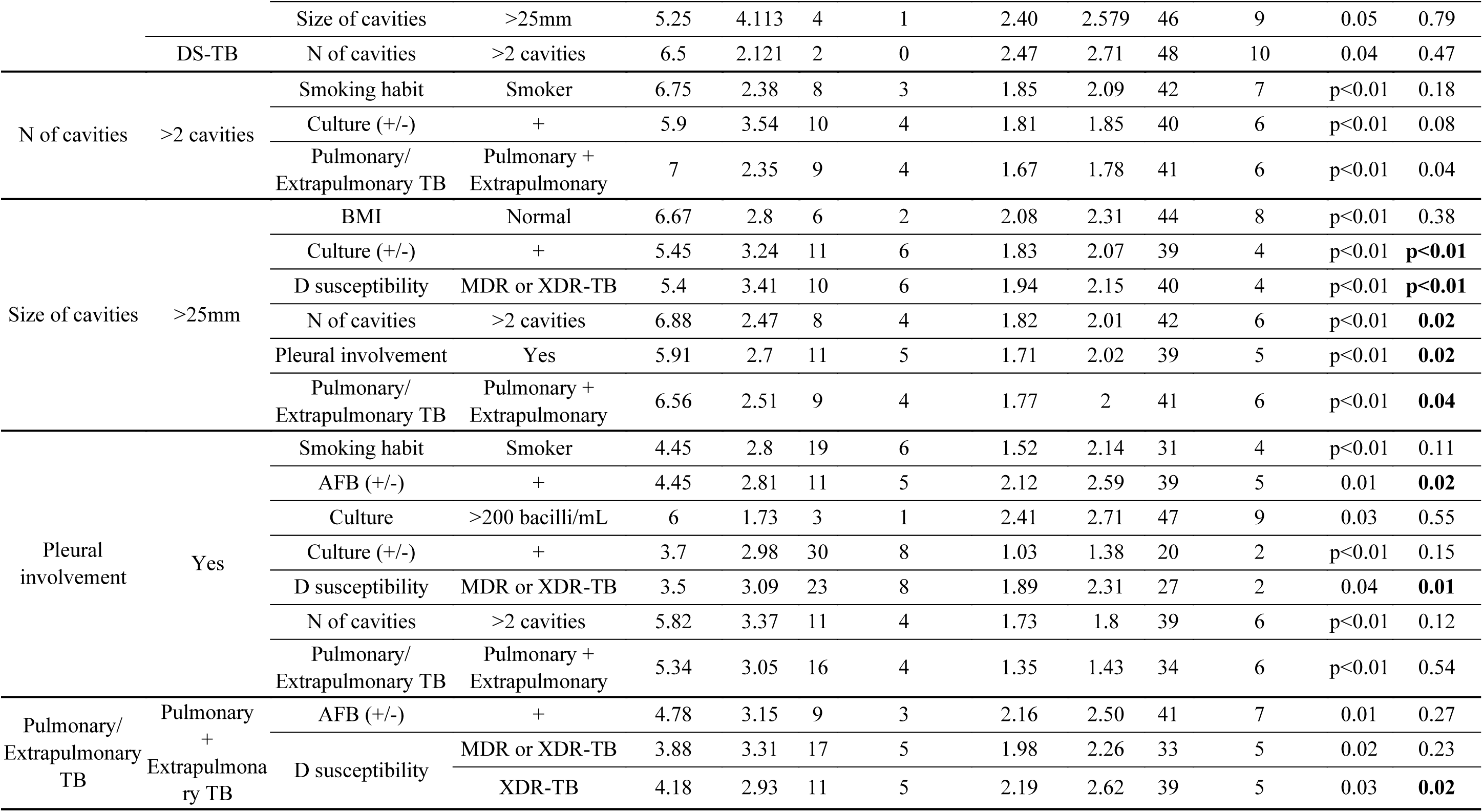
Values of the index of signs of activity at s₀ in patients presenting both variable 1 and variable 2 versus patients presenting either variable 1 or variable 2 or neither, along with the corresponding p-value. *The p-value for the final outcome of treatment failure or death in patients from the first group and the second group is also provided. p-values ≤ 0.05 are highlighted in bold*.

This motivated us to identify general index thresholds at BL for activity signs that could predict the likelihood of cure/treatment completion versus treatment failure/death. Patients were grouped based on their activity index at BL and the frequency of their final outcomes. The probability of these outcomes was then calculated according to the index value (Figure 3a). A total of 80% of patients with a final outcome of treatment failure or death had an index at BL ≥ 2. Of the 10 patients with this outcome, 8 (80%) had an index ≥ 2; and the 80% of patients with a final outcome of cure/treatment completion had an index < 3. Of the 36 patients with this outcome, 28 (80%) had an index < 3 (Figure 3b).

**Figure 3.**
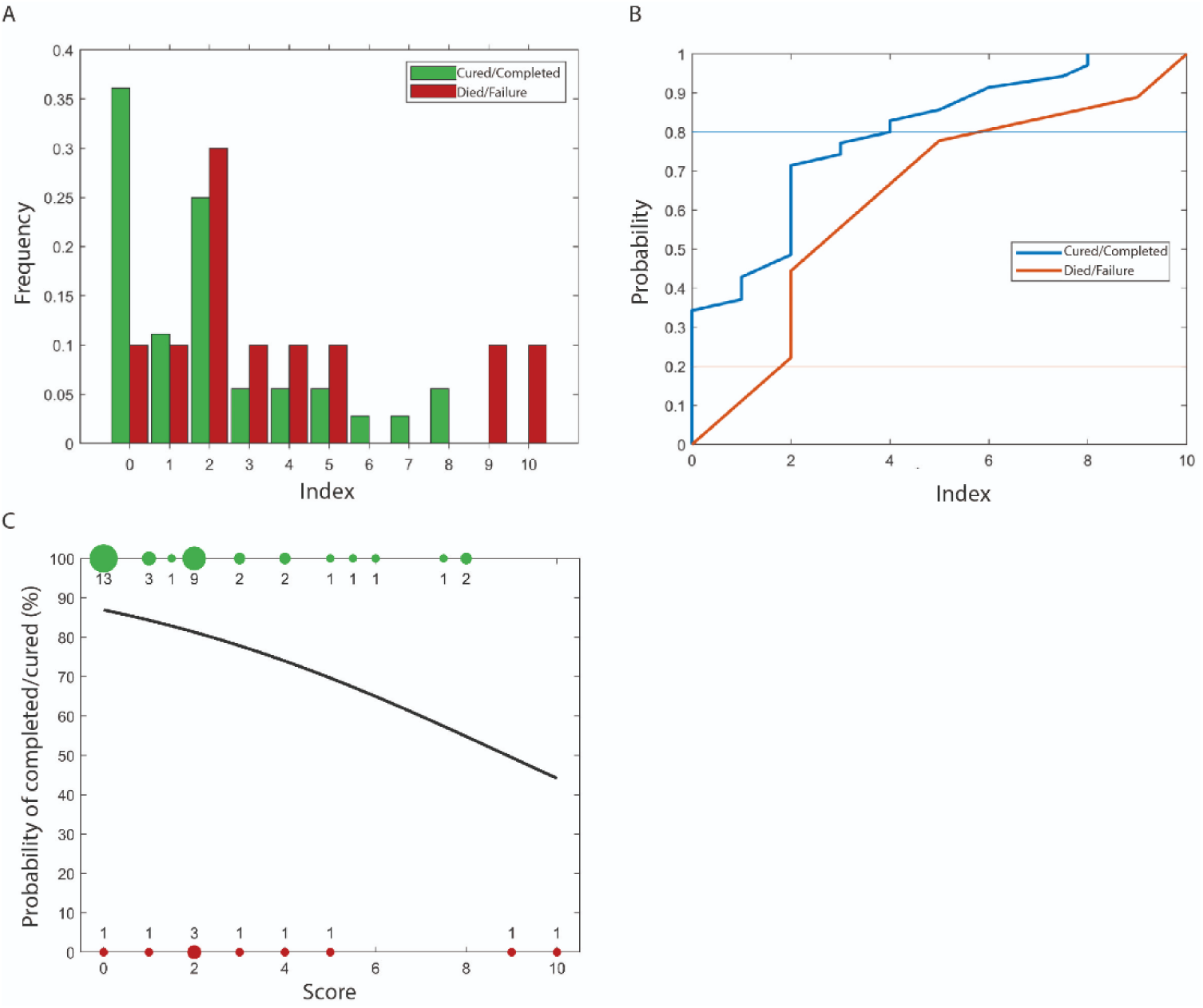
**Panel A**: Frequency of index scores at s₀ for signs of activity, depending on the patient’s final status. **Panel B:** probability of presenting the final outcomes—cured/completed and failure/death— depending on the index score. **Panel C**: Logistic regression showing the probability of final outcomes (treatment completed/cured) relative to the index score.

Logistic regression results for predicting the probability of cure/treatment completion are shown in Figure 2c. The probability of cure was 90% with an index of 0, decreasing to 50% with an index of 8. It is important to note that the sample size was small, resulting in wide confidence intervals. A larger sample could provide more precise probability estimates and refine the model parameters. Nonetheless, the analysis showed a clear trend: the probability of cure decreased as the index increased.

Based on the studied cohort, it appears that the threshold for distinguishing between a favourable and an unfavourable final outcome lies between an index of 2 and 3 for activity signs at BL. A larger sample size would be necessary to confirm this trend. The probability of experiencing a final outcome of treatment failure or death was found to be proportional to the baseline activity index value.

## 4. Discussion

This study designed and optimized a radiological index by testing it on a small, homogeneous cohort of individuals with pulmonary TB and further validated in a larger independent cohort. Our results demonstrate its utility in quantifying the severity of active TB on a baseline CXR and tracking disease evolution over time. Radiological signs of inactive TB remained stable during F-U. A correlation was established between the index values for active TB signs on the baseline CXR and various clinical and microbiological variables, both individually and in pairs. Additionally, the BL activity index showed prognostic value for unfavourable outcomes, indicating its potential for adjusting treatment in high-risk patients.

CXR has been a cornerstone in the diagnosis and follow-up of pulmonary TB patients since the late 19th century, forming a critical part of diagnostic algorithms. If the severity of pulmonary involvement at disease diagnosis determines the patient’s final outcome post-treatment, and if CXR can accurately assess this severity, then it should also predict final outcomes. Effective observation and interpretation of radiological findings could aid in predicting worse outcomes, guiding therapeutic strategies for patients at risk. This is not the first index reported. Different clinical indices for assessing pulmonary TB have been described in the literature, often with simplified radiological assessments based solely on the presence or absence of specific findings. For example, Bock et al. and Mylotte et al. created indices combining clinical and radiological data to identify patients requiring isolation upon hospital admission, focusing primarily on upper lobe infiltrates or cavitation [17, 26]. Other studies, including those by Wisnivesky et al., Racokzy et al., and Lagrange et al., developed similar clinical-radiological indices during the 1990s and early 2000s [15, 20, 21, 23]. Subsequent efforts aimed to create predictive indices for pulmonary TB diagnosis in symptomatic, smear-negative patients in emergency settings. Soto et al. and Solari et al. proposed clinical-radiological indices that included apical infiltrates and miliary involvement, validated later for predictive accuracy [12–14, 24]. In 2010, Ralph et al. developed a numerical radiological index, the *Timika X-ray score*, to assess severity in smear-positive TB patients. It evaluated nodules, consolidations, cavitations, bronchial lesions, fibrosis, lymphadenopathy, and pleural effusion, dividing the lung into three zones for assessment[4]. Pefura Yone et al. later proposed a simplified radiological index based on Ralph’s work, adding a weighting for cavity presence [16]. Contributions of the index developed in this study compared to existing indices is presented in Table 5.

**Table 5.** Comparison of Parameters Assessed by the RTBES and Other Existing Indices.

| Index & its qualities | Simplicity | Interobserver Concordance | Severity Assessment | F-U | Parameter for Final Outcome Measurement | Predictive Value for Patient's Final Outcome |
| --- | --- | --- | --- | --- | --- | --- |
| *SA-BCN score* | +++ | 83-96% | Yes | Yes | Final status at the end of treatment | Yes |
| *Timika score*<br>Ralph et al | + | 31-81%<br>Depending of the evaluated variables | Yes | Not included | Microscopy at two months of treatment | Yes |
| *Timika score*<br>Krishnamoorthy et al |  |  |  |  | Final status at the end of treatment | No |
| *Timika score modified*<br>Kriel et al | + | 86% | Yes | Not included | Microscopy at two months of treatment | Yes |
|  |  |  |  |  | Final status at the end of treatment | No |
| **SCS**<br>Pefura-Yone et al | + | Evaluation done by a single radiologist | Yes | Not included | Microscopy at two months of treatment | Yes |

The development of the RTBES successfully met the initial design criteria: simplicity, reliability, accuracy, and numerical quantification of TB findings. Additionally, it was designed to be adaptable for automated analysis. The index is presented with concise instructions and a graphical format, contrasting with most literature indices, which are often presented as checklists. A graphical format for the RTBES was created, with a schematic representation of lungs, pleural spaces, and lymph nodes, making it simple for radiologists and non-radiologists to use, even by non-medical personnel. The scoring criteria for active and inactive TB are straightforward, with concise instructions for completion.

Unlike other indices, the RTBES incorporates a broader range of radiological TB presentations, including pulmonary infiltrates, cavities, bronchogenic dissemination, and miliary patterns. Its scoring method is simpler yet more precise than a visual assessment of affected lung percentage. Once adjusted, the index accurately evaluated TB severity at baseline and its progression over time. As observed in the first part of the study, there was a slight decrease in the activity index at the first F-U, with a more pronounced reduction at the second follow-up, reflecting the gradual improvement typical in TB treatment, where clinical recovery often precedes radiological improvement. Ralph et al. similarly demonstrated a decrease in their index over time [4].

The RTBES was developed using data from a homogeneous cohort with high HIV prevalence and validated in a geographically distinct cohort with a higher prevalence of drug-resistant TB. This approach allowed for testing the index across a spectrum of radiological presentations, enhancing its reliability. Both cohorts consisted of outpatients. This is consistent with other indices validated in outpatient populations, often with lower HIV co-infection and drug-resistant TB prevalence [4, 9, 16]. However, it is important to consider that when immune status is preserved, radiological findings are similar to those in non-HIV patients while with severe immunosuppression, up to 10-40% of CXRs may appear normal, despite significant pulmonary disease [27, 28]. In these cases, the RTBES might not be applicable.

In terms of interobserver agreement, the RTBES demonstrated comparable reliability to existing indices like the Timika score and its modified versions [29–31]. As with other indices, it accurately assessed initial TB severity, a finding supported by Thiel et al., who confirmed the predictive utility of the Timika score [29, 32]. Interobserver variability was assessed using two radiologists – one experienced, the other a resident. Literature review shows diverse readers in similar studies, ranging from radiologists to non-radiologists, trained or untrained in CXR interpretation. In clinical practice, outpatient radiological evaluations are frequently conducted by non-radiologists, potentially increasing variability. Targeted training on TB-specific radiological findings could mitigate this variability.

The predictive value of the RTBES for final outcomes underscores its potential in clinical practice. It offers an opportunity to identify individuals at risk who might benefit from treatment adjustments, including the incorporation of active host-directed therapies to mitigate extensive lung damage, a potential benefit proposed in recent years [33–35]. Further studies incorporating the RTBES to quantify radiological extent and its evolution during TB management in randomised clinical trials could provide additional insights while also serving as external validation for the tool.

Finally, although Artificial Intelligence (AI) systems have been developed for pulmonary and TB diagnosis, current systems are not yet certified for evolutionary assessment or outcome prediction [36, 37]. Applying AI tools to the RTBES could automate its use and enhance its predictive capabilities. When properly trained and validated, AI algorithms can provide highly consistent segmentations and, once deployed, rapidly process large numbers of images while harmonizing scoring across different personnel and institutions. Indeed, studies have demonstrated that AI tools can successfully assist physicians in detecting CXR abnormalities for certain findings, even though these algorithms may miss subtle clinical or contextual nuances that a trained radiologist would detect [38, 39]. In low-resource settings, while AI might help overcome radiologist shortages, it can also be too expensive or challenging to deploy on-site, and many implementation challenges remain to be addressed [40]. Overall, developing or adapting an AI-based tool for the RTBES holds significant promise for automating its use and enhancing its predictive performance.

In conclusion, the RTBES provides an effective means to quantify the severity of radiological signs of TB on baseline CXR and to monitor and predict the progression and regression of the disease throughout treatment. The RTBES stands out as a simple, easy-to-use, reproducible, and precise tool with minimal interobserver variability. Notably, certain clinical and microbiological parameters are associated with RTBES values for signs of activity at BL. Importantly, our findings indicate that the probability of an unfavourable final outcome is directly proportional to the RTBES value for signs of activity at BL. Therefore, the RTBES is a valuable instrument in clinical practice for risk stratification and management of patients with active TB.

## Data Availability

All data produced in the present work are contained in the manuscript

## 6. Authorship statement

JB and CV conceived the score and the work. PC, RP, MT, JB and CV made substantial contributions to the design of the work. PC, GR, MC, NM, AR, AG and SV made substantial contributions to the acquisition and/or analysis of data for the work. PC, MC, RP, MT, JB and CV interpreted the data. PC, MC, LA and CV drafted the article. All authors revised the manuscript draft for important intellectual content, gave final approval of the version to be published; and agreed to be accountable for all aspects of the work, in ensuring that questions related to the accuracy or integrity of any part of the work are appropriately investigated and resolved.

## 7. Acknowledgements

We used ChatGPT (OpenAI, ChatGPT, version 4) to help improve the readability and clarity of this manuscript. The text was reviewed and edited by the authors, who assume full responsibility for the final content.

## Procedure for the Quantitative Assessment of Tuberculosis Evolution Using the Radiological Index RUTI-TB Evolution Score (RTBES)

Authors: Patrícia Cuadras, Gerard Rafart, Martí Català, Lilibeth Arias, Ricard Pérez, Montserrat Tenesa, Jordi Bechini^&^ and Cristina Vilaplana^&^.

^&^These authors contributed equally to this work

Affiliation: Unitat de Tuberculosi Experimental, Germans Trias i Pujol Research Institute and Hospital (IGTP-HUGTIP), Can Ruti Campus, Ctra. del Canyet, S/N, 08916 Badalona, Catalonia, Spain

Contact author: Cristina Vilaplana

## 1. Procedures

A suitably qualified professional will analyse each patient’s Chest X-Ray (CXR) and follow the specified procedure to calculate the RUTI-TB Evolution Score (RTBES). A dedicated form available at the end of this document will be printed and retained for each patient throughout the clinical trial or routine clinical visits.

The procedures described by the RTBES authors are as follows.

### Before starting

1- If the technical quality of the CXR to be evaluated is suboptimal, do not use it.
2- Ensure that the CXR to be evaluated are performed with the patient in a standing position.

### Procedure

The RTBES requires evaluating multiple CXRs at specified intervals, either based on clinical follow-up (FU) or the study protocol (e.g., baseline, week 8, and week 24, with an additional CXR at the end of treatment for MDR patients). Further follow-up CXRs may be included if clinically appropriate.

1- Complete the patient form.
2- For each timepoint, use the RTBES form to document as described in the subsequent sections.

a. Signs of activity
b. Signs of non-activity
3- Start with the qualitative analysis: Consider both the right and left lungs. From the right hilum, draw an imaginary horizontal line to divide each lung into two equal parts: superior and inferior (50%-50%) (Figure 1). Based on the previously mentioned division of the lungs, four quadrants will be generated in which the affected areas for each item must be indicated using the template provided at the end of this document.

**Figure 1:**
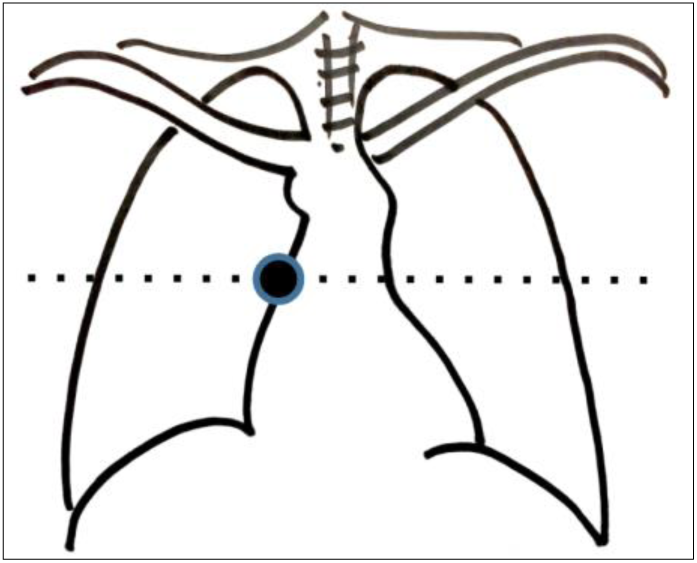
Lung for qualitative analysis. From the right hilum (black dot) a horizontal dotted line divides each lung in two equal parts, superior and inferior.
4- Proceed with the Quantitative Analysis: Assign a score to each identified affected area in accordance with the rules outlined in Tables 1 (Signs of activity) and 2 (Signs of non-activity).

**Table 1:**
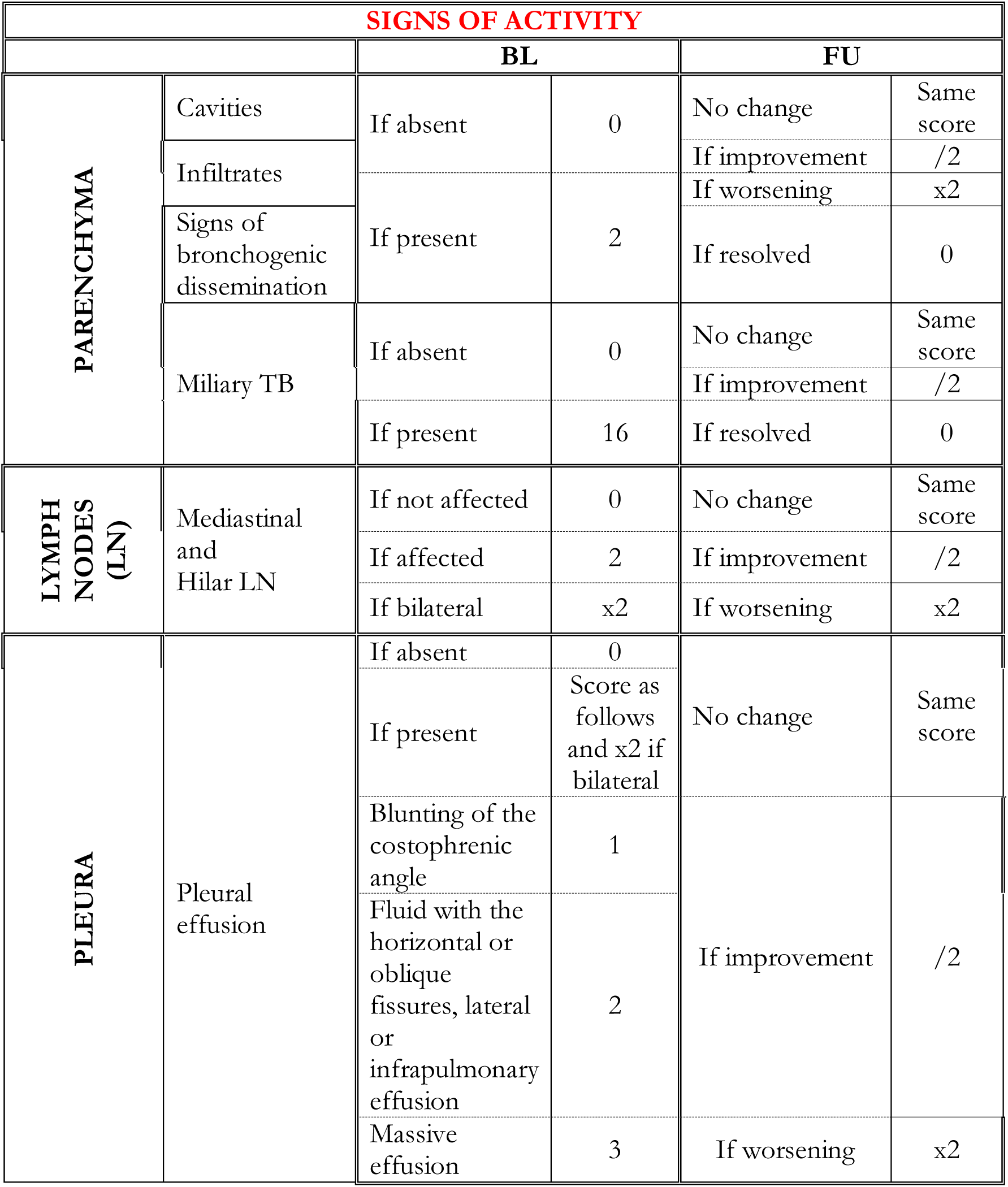
Signs of activity.

**Table 2:** Sings of chronicity (non-active X-ray findings)

| **NON-ACTIVE X-RAY FINDINGS** |  |  |  |  |  |
| --- | --- | --- | --- | --- | --- |
|  |  | **BL** |  | **FU** |  |
| **PARENCHYMA** | Cicatricial lesions | If absent | 0 | No change | Same score |
|  | Calcified granulomas |  |  |  |  |
|  | Atelectasis |  |  |  |  |
|  | Bronchiectasis | If present | 2 | If worsening | x2 |
|  | Destructive pattern |  |  |  |  |
| **LN CALCIFICATION** | Mediastinal and Hilar LN | If not affected | 0 | No change | Same score |
|  |  | If affected | 2 | If improvement | /2 |
|  |  | If bilateral | x2 | If worsening | x2 |
| **PLEURA** | Thickening, calcification | If absent | 0 | If absent | 0 |
|  |  | If present | 2 | If present | 2 |
|  |  | If bilateral | x2 | If bilateral | x2 |

For each assessment timepoint, calculate and document both individual subscores (Activity X-ray signs subscore and Non-Active X-ray signs subscore), as well as the total score (the sum of these two subscores).

Record all values in the medical record or the CRF.

Use the total score for an initial assessment of the patient and rely on the Activity X-ray signs subscore to gauge radiological improvement or worsening over time.

## 2. Template

Template to be used for assessing the TB-related Activity and Non-activity Signs in the CXR and calculate the RTBES can be found here below.

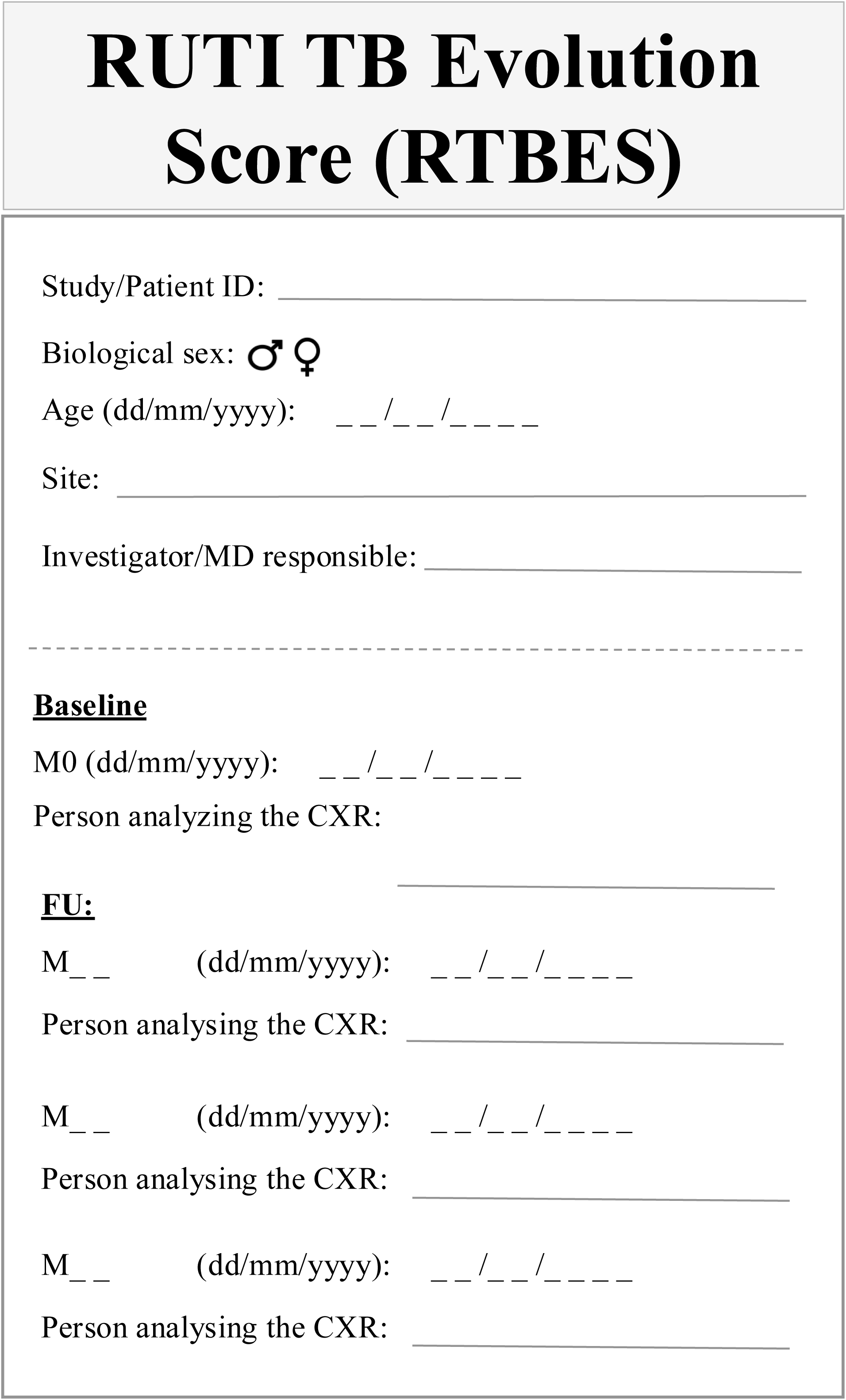

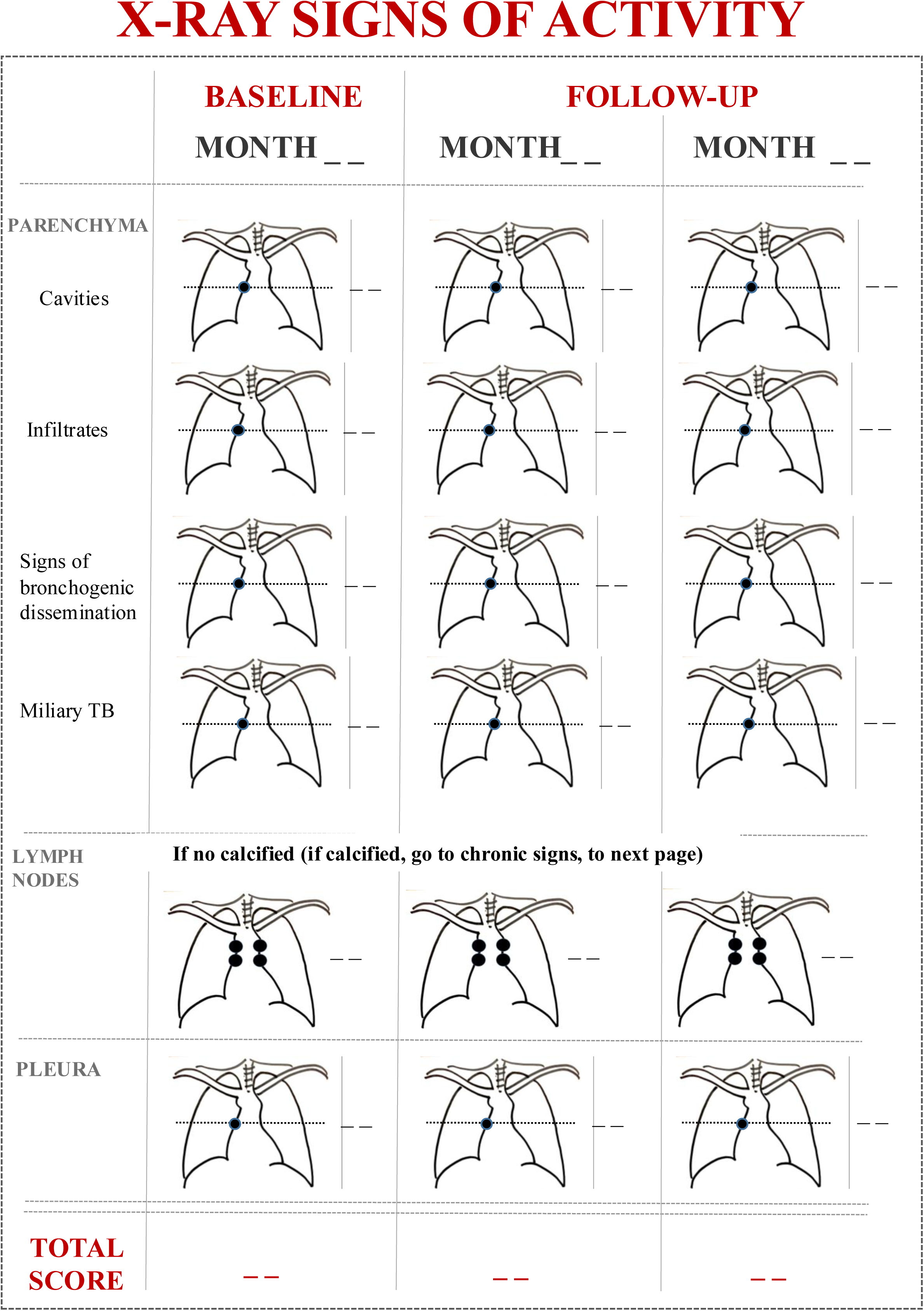

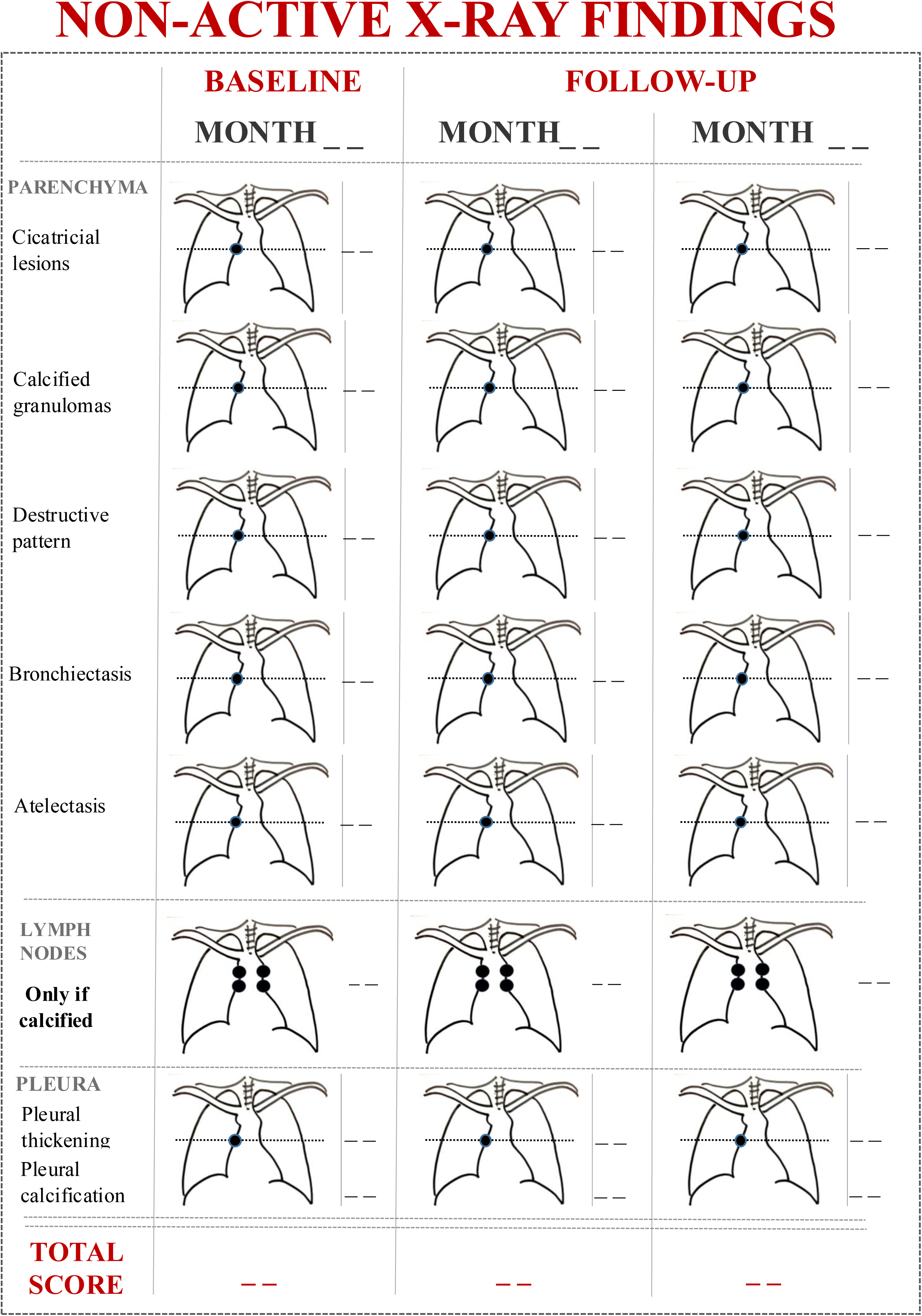

